# Comparison of structural MRI brain measures between 1.5T and 3T: data from the Lothian Birth Cohort 1936

**DOI:** 10.1101/2021.04.23.21256000

**Authors:** Colin R. Buchanan, Susana Muñoz Maniega, Maria C. Valdés Hernández, Lucia Ballerini, Gayle Barclay, Adele M. Taylor, Tom C. Russ, Elliot M. Tucker-Drob, Joanna M. Wardlaw, Ian J. Deary, Mark E. Bastin, Simon R. Cox

**Affiliations:** Lothian Birth Cohorts group, The University of Edinburgh, UK; Department of Psychology, The University of Edinburgh, UK; Scottish Imaging Network, A Platform for Scientific Excellence (SINAPSE) Collaboration, Edinburgh, UK; Centre for Clinical Brain Sciences, The University of Edinburgh, UK; Alzheimer Scotland Dementia Research Centre, The University of Edinburgh, UK; Department of Psychology, University of Texas, Austin, Texas, USA

**Keywords:** structural MRI, diffusion MRI, multi-site, reliability, brain, connectome

## Abstract

Multi-scanner MRI studies are reliant on understanding the apparent differences in imaging measures between different scanners. We provide a comprehensive analysis of T1-weighted and diffusion MRI (dMRI) structural brain measures between a 1.5T GE Signa Horizon HDx and a 3T Siemens Magnetom Prisma using 91 community-dwelling older participants (aged 82 years). Although we found considerable differences in absolute measurements (global tissue volumes were measured as ∼6—11% higher and fractional anisotropy was 33% higher at 3T than at 1.5T), between-scanner consistency was good to excellent for global volumetric and dMRI measures (intraclass correlation coefficient [ICC] range: 0.612—0.993) and fair to good for 68 cortical regions (FreeSurfer) and cortical surface measures (mean ICC: 0.504—0.763). Between-scanner consistency was fair for dMRI measures of 12 major white matter tracts (mean ICC: 0.475—0.564), and the general factors of these tracts provided excellent consistency (ICC ≥ 0.769). Whole-brain structural networks provided good to excellent consistency for global metrics (ICC ≥ 0.612). Although consistency was poor for individual network connections (mean ICCs: 0.275 – 0.280), this was driven by a large difference in network sparsity (0.599 versus 0.334), and consistency was improved when comparing only the connections present in every participant (mean ICCs: 0.533—0.647). Regression-based k-fold cross-validation showed that, particularly for global volumes, between-scanner differences could be largely eliminated (R^2^ range 0.615—0.991). We conclude that low granularity measures of brain structure can be reliably matched between the scanners tested, but caution is warranted when combining high granularity information from different scanners.

## Introduction

Understanding how estimates of brain structure vary across different MRI hardware and field strengths is an important aspect of neuroimaging research since knowledge of between-scanner differences and the means to match measures between scanners is an essential prerequisite for multi-site analyses or multi-scanner longitudinal studies. Cross-scanner comparisons of brain measures across scanner hardware is useful in multiple settings, including hardware replacement, relocation in ongoing research studies or pooling of multi-site data (Kruggel, Turner, & Muftuler, 2010). An increase in the use of 3T scanners is driven in part by the potential to increase tissue contrast and reduce background noise (thereby increasing the signal-to-noise ratio and contrast-to-noise ratio), the ability to acquire higher resolution scans more quickly, to acquire higher b-values and thinner slices in diffusion MRI (dMRI) and advanced methods such as neurite orientation dispersion and density imaging (Zhang, Schneider, Wheeler-Kingshott, & Alexander, 2012), and to potentially increase diagnostic accuracy (Fushimi et al., 2007; Schmitz, Aschoff, Hoffmann, & Grön, 2005; Tanenbaum, 2005; Wardlaw et al., 2012). In clinical practice, although higher field strength MRI may improve image quality and diagnostic accuracy, the theoretical doubling of the signal-to-noise ratio in practice was only 25%, though 3T appeared to outperform 1.5T technology in research settings (Wardlaw et al., 2012). Although the possibility of combining MRI data points from different scanner hardware is appealing, this is challenging because scanner-dependent geometric distortions and differences in tissue contrast can be problematic (Gunter et al., 2009).

When considering the potential for two different field strengths to yield different estimates of the same brain measurements, interpretation should be tempered by the finding that even same-scanner measurements, taken twice or more over short periods, are not perfectly reliable. Same-scanner test-retest studies of the same subjects report agreement as low as 0.8 in terms of the intraclass correlation coefficient (ICC) for global volumetric and water diffusion measures (Iscan et al., 2015; Luque Laguna et al., 2020; Melzer et al., 2020). Agreement is generally lower for specific regional volumetric measures, where cortical thickness ICCs > ∼0.50 have been reported (Madan & Kensinger, 2017) and ICCs > ∼0.8 even when both acquisitions were taken in the same session (Liem et al., 2015). Similarly, tract-or region-specific diffusion measures have been reported with ICCs < 0.54 (Luque Laguna et al., 2020), ICCs > 0.72 (Boekel, Forstmann, & Keuken, 2017) and coefficient of variation (CoV) < 10% (Clayden, Storkey, Maniega, & Bastin, 2009). This trend is echoed by structural connectomic measures, in which global network properties had more reliable (but still imperfect) test-retest agreement (ICCs > ∼0.6), in contrast to lower reliabilities (ICCs > 0.46) of regional/nodal properties (Buchanan, Pernet, Gorgolewski, Storkey, & Bastin, 2014; Cheng et al., 2012).

In this context, the extant cross-field comparison studies which compare brain MRI measures between 1.5T and 3T indicate that agreement, in small samples of generally younger participants, may not be substantially lower than same-scanner test-retest findings. For example, between-field-strength differences have been reported as < 10% for brain and tissue volumes (Heinen et al., 2016) and ∼10% for subcortical measurements (Chu, Hurwitz, Tauhid, & Bakshi, 2017). White matter (WM) diffusion measures have been reported with CoV < 7.5% (Grech-Sollars et al., 2015). Regional cortical measures have also shown relatively acceptable to good reliability across scanner manufacturer and field strength in some samples (Han et al., 2006; Pfefferbaum, Rohlfing, Rosenbloom, & Sullivan, 2012; Reuter, Schmansky, Rosas, & Fischl, 2012; Wonderlick et al., 2009), whereas others report discrepant results (Gronenschild et al., 2012; Morey et al., 2010; Srinivasan et al., 2020). In some studies, global properties such as volume and cortical thickness are generally larger at higher field strengths (Chu et al., 2017; Han et al., 2006; Heinen et al., 2016; Pfefferbaum et al., 2012), though West et al. (2013) reported that whereas grey matter (GM) and cerebrospinal fluid (CSF) volume appeared higher at 3T, WM volumes were lower.

However, these studies have typically been conducted using modest sample sizes (often N ≤ 20, but see Pfefferbaum et al., 2012 and Srinivasan et al., 2020) and among adults almost exclusively younger than 65 years old. Brains that, on average, exhibit greater degeneration are ‘further’ from the average atlases upon which some neuroimaging pipelines rely, and older participants have a greater array of physical limitations which are a barrier to achieving artefact-free imaging data during extended scanning sessions (e.g. arthritis). The wider variability in structural brain measures among individuals who are at greatest risk of cognitive decline and a range of age-related diseases and disorders might hamper the generalisability of findings from younger groups. Moreover, the low sample sizes mean that any statistical analyses aimed at identifying meaningful differences between conditions are likely to be substantially underpowered, providing speculative estimates of the comparability of data across field strengths. Finally, only a single, or a small subset, of MRI-derived phenotypes has been considered at once. Such factors fundamentally complicate the meaningful synthesis of extant data for assessing the likely cross-scanner impact on structural and diffusion measures in older participants.

To address these gaps in the literature, the current study assesses an array of T1-weighted and dMRI imaging variables using 91 participants, aged 82 years, scanned at both 1.5T and 3T. Between-scanner comparison of imaging variables was performed at several levels: overall brain and tissue volumes; regional cortical and subcortical GM volumes; cortical volume, surface area and thickness; dMRI measures in global WM, dMRI measures in 12 WM tracts; and whole-brain structural networks. Additionally, we used 10-fold cross validation to test prediction of ‘unseen’ 1.5T values from 3T data using linear regression.

## Materials and methods

### Participants

Data were drawn from the Lothian Birth Cohort 1936 (LBC1936), an on-going study on the influences on cognitive ageing from age 11 into the 8th and 9th decades of life (Deary et al., 2007; Deary, Gow, Pattie, & Starr, 2012; Taylor, Pattie, & Deary, 2018). Structural imaging including dMRI has been performed on the same well-maintained 1.5T scanner at all imaging waves (Wardlaw et al., 2011). A subset of participants were also imaged at 3T. This was motivated by the intention to safeguard against potential unexpected breakdown of the scanner, or facility relocation, and an incentive to use modern MRI acquisitions, reducing participant burden in terms of comfort and duration for those who are becoming increasingly frail and less able to lie still in a scanner for the hour-long 1.5T acquisition. A total of 105 (60, 57.1% male) community-dwelling participants in the Lothian area were therefore recruited from the fifth wave of the LBC1936. Prior to undergoing either scan, participants who had indicated they would undergo the standard 1.5T session were invited to also undergo a 3T imaging session – then, following a successful 1.5T scan they were booked for 3T imaging. Participants were recruited on a first-come, first-served basis and 3T imaging ended after 105 participants had successfully completed both scans. The mean interval between scans was 71.9 (SD = 16.6; range 28-111) days. At the time of the 1.5T scan, participants had a mean age of 82.0 (SD = 0.3) years. Written informed consent was obtained from each participant under protocols approved by the Lothian (REC 07/MRE00/58) and Scottish Multicentre (MREC/01/0/56) Research Ethics Committees.

### MRI acquisition

MRI acquisition parameters at 1.5T have been described previously (Wardlaw et al., 2011). All participants underwent brain MRI on the same 1.5T GE Signa Horizon HDx clinical scanner (General Electric, Milwaukee, WI, USA) with a manufacturer supplied 8-channel phased-array head coil. High resolution 3D T1-weighted inversion-recovery prepared, fast spoiled gradient-echo volumes were acquired in the coronal plane with 180 contiguous 1.3 mm thick slices resulting in voxel dimensions of 1 × 1 × 1.3 mm. For the dMRI protocol, single-shot spin-echo echo-planar (EP) diffusion-weighted whole-brain volumes (*b* = 1000 s mm^− 2^) were acquired in 64 non-collinear directions, along with seven T2-weighted volumes (*b* = 0 s mm^− 2^). Seventy-two contiguous axial 2 mm thick slices were acquired resulting in 2 mm isotropic voxels.

The same 105 participants had a brain MRI on a 3T Siemens Magnetom Prisma (Siemens Healthcare Gmbh, Erlangen, Germany) using a 32-channel matrix phase array head coil. High resolution 3D T_1_-weighted MPRAGE volumes (TR = 2520 ms, TE = 4.37 ms, TI = 1270 ms, flip angle = 7°, FOV = 256 × 208 mm, acquisition time = 3 min, 12 s) were acquired in the coronal plane with 224 contiguous 1 mm thick slices resulting in 1 mm isotropic voxels. The multi-shell dMRI protocol employed a single-shot spin-echo EP diffusion-weighted sequence which acquired 14 b = 0 s mm^-2^, 3 b = 200 s mm^-2^, 6 b = 500 s mm^-2^, 64 b = 1000 s mm^-2^ and 64 b = 2000 s mm^-2^ whole-brain volumes (TR = 4300 ms, TE = 74 ms, FOV = 256 mm, acquisition matrix 128 ×128, total acquisition time = 11 min, 12 s). Seventy-four contiguous axial 2 mm thick slices were acquired resulting in 2 mm isotropic voxels. A reverse phase encoding EP dataset with 6 b = 0 s mm^-2^ whole brain volumes was also collected for subsequent EP susceptibility distortion correction using the same acquisition parameters as the main dMRI protocol. Two participants were excluded from T1-weighted analyses and five were excluded from dMRI analyses due to incomplete or missing scans at 1.5T.

### T1-weighted processing

Volumetric segmentation and cortical reconstruction were performed with the FreeSurfer image analysis suite (http://surfer.nmr.mgh.harvard.edu) version 6.0.0. The Desikan-Killiany atlas delineated 34 cortical structures per hemisphere (Desikan et al., 2006; Fischl, Van Der Kouwe, et al., 2004). Subcortical segmentation was applied to obtain eight GM structures per hemisphere: accumbens area, amygdala, caudate, hippocampus, pallidum, putamen, thalamus and ventral diencapahlon (Fischl et al., 2002; Fischl, Salat, et al., 2004). Grey and white tissue matter masks were obtained. Total CSF volume was computed as the sum of the volumes of the ventricular system, nonventricular CSF and choroid plexus.

We applied FreeSurfer using the default parameters and opted not to undertake any manual editing so as not to introduce any rater-specific bias into the comparison between scanners. The outputs of FreeSurfer were manually quality checked (QC by CB and SC) to exclude participants with severe motion artefact or gross segmentation errors. Participants with brain lesions or suspected stroke (N=8) were not excluded from our sample as we intended to compare between-scanner segmentation with representative data of our older age sample. Eight participants failed QC at 1.5T, seven participants failed at 3T, and in total 91 participants passed QC at both 1.5 and 3T. For each region delineated by the FreeSurfer procedure the total volume was recorded based on volume in T1-weighted space. Cortical surface analyses were performed using the SurfStat MATLAB toolbox (http://www.math.mcgill.ca/keith/surfstat). Surfaces were aligned vertex-wise into a common space (the FreeSurfer average template) and spatially smoothed at 20⍰mm full width at half maximum (FWHM), allowing sample-wide analyses of volume, area and thickness across the cortex. In a supplementary analysis to test the efficacy of smoothing, we also computed the same measures over a range of smoothing widths from 0 to 25 mm.

### Diffusion MRI processing and tractography

The 1.5T dMRI raw data was read and converted from DICOM to NIfTI-1 format using TractoR v2.6.2 (http://www.tractor-mri.org.uk; Clayden et al., 2011). Using tools freely available in the FSL toolkit v4.1.9 (FMRIB, Oxford University: http://www.fmrib.ox.ac.uk; Smith et al., 2004), data underwent brain extraction (Smith, 2002) performed on the T2-weighted EP volumes acquired along with the dMRI data. The brain mask was applied to all volumes after correcting for systematic eddy-current induced imaging distortions and bulk patient motion using affine registration to the first T2-weighted EP volume of each subject with ‘eddy_correct’ (Jenkinson & Smith, 2001). Due to the longitudinal character of the LBC1936 study, the dMRI processing protocol at 1.5T has remained unchanged since the first LBC1936 imaging wave in 2007.

We determined that alternative processing was required for the 3T dMRI data due to greater susceptibility-induced distortions at this field strength. This data was read and converted from DICOM to NIfTI-1 format using TractoR v 3.3.1, masked using FSL’s brain extraction tool (Smith, 2002) and corrected for susceptibility and eddy current induced distortions using ‘topup’ and ‘eddy’ from FSL version 5.0.9 (Andersson, Skare, & Ashburner, 2003; Andersson & Sotiropoulos, 2016). Additionally, to test the impact of different pre-processing pipelines between scanners we also applied the 1.5T pipeline (Tractor v2.6.1 and FSL v4.1.9) using 3T data for 10 subjects.

For all dMRI volumes, diffusion tensors were fitted at each voxel using FSL’s ‘dtifit’ and water diffusion measures were estimated for axial (AD), radial (RD) and mean (MD) diffusivity, which measure magnitudes of molecular water diffusion. Fractional anisotropy (FA) was also computed, which measures the degree of anisotropic diffusion per voxel (Pierpaoli & Basser, 1996). All diffusion measures and tractography were computed in diffusion space. Mean values of the four dMRI measures were estimated in cerebral WM (using the WM mask obtained from FreeSurfer aligned to diffusion space). In a supplementary analysis we also computed the same measures across the whole brain (using the mask obtained from the T2-weighted EP volume).

Whole-brain tractography was performed using an established probabilistic algorithm (BEDPOSTX/ProbtrackX; Behrens et al., 2007, 2003). Probability density functions, which describe the uncertainty in the principal directions of diffusion, were computed with a two-fibre model per voxel (Behrens et al., 2007). Streamlines were then constructed by sampling from these distributions during tracking with a fixed step size of 0.5 mm between successive points.

Analysis of 12 major WM tracts was performed using probabilistic neighbourhood tractography (PNT; Clayden et al., 2011). These tracts were the genu and the splenium of the corpus callosum, and bilateral arcuate fasciculi, anterior thalamic radiations (ATR), frontal projection of the cingulum bundles, uncinate and inferior longitudinal fasciculi (ILF). Although the ventral cingulum bundles were also computed these were excluded from analysis because they have previously been deemed unreliable. All tracts were visually quality checked (SMM) and exclusions were made on a tract basis. Probability maps showing the density of streamlines in each tract were computed across participants for those who had validated tract data. The weighted mean values of MD and FA were computed per tract. Consistent with prior work in the full LBC1936 sample (Ritchie et al., 2015), we also extracted general factors (gMD and gFA) by performing principal component analysis on the tract data and extracting the first unrotated principal component.

### Network analysis

Whole-brain structural networks were computed for 79 participants (who had passed both T1 and dMRI QC at both field strengths). Networks were constructed using 85 neuroanatomical regions (the 84 GM regions described above plus the brain stem) and probabilistic tractography resulting in 85×85 networks (Buchanan et al., 2020). Networks were computed for both MD and FA by computing the mean value of each measure in all voxels along the interconnecting streamlines between a pair of regions. We applied network thresholding using consistency-thresholding (Roberts, Perry, Roberts, Mitchell, & Breakspear, 2017) to remove some proportion of putatively spurious connections across subjects at a threshold level retaining the top 30% most consistent connections that was previously determined from a large single-scanner study (Buchanan et al., 2020). To obtain a representative estimate of between-scanner agreement, thresholding was applied separately for both field strengths (resulting in non-identical sets of connections). Three common global graph-theoretic metrics were computed using weighted measures (Rubinov & Sporns, 2010): mean edge weight (mean of all edge weights per subject), global network efficiency (a measure of integration) and network clustering coefficient (a measure of segregation).

### Statistical analysis

Between-scanner comparison of imaging variables was performed at several levels: global (overall tissue volumes, dMRI measures in WM, global network metrics), regional (GM regions, major WM tracts) and sub-regional level (cortical surface vertex analysis, networks connections). Imaging variables were paired between scanners and we computed both the between-scanner difference and ICC to assess agreement. For a paired set of subject-specific measures, 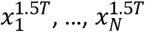 and 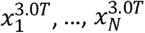, the average *between-scanner difference* was computed,

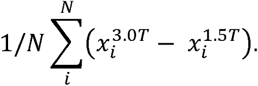

Similarly, the mean between-scanner difference expressed at percent change from the 1.5T values was computed,

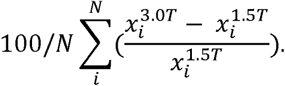

We computed ICC(3,1) using two-way mixed single measures (Shrout & Fleiss, 1979) using *consistency* of measurements between sessions. The ICC was originally formulated for assessing multiple raters in measuring the same quantity. We computed ICC between scanners for each imaging measure by measuring the 1.5T subject-specific values against the same values at 3T. For ICC scores we adopted the four level rating from (Cicchetti, 1994): *poor* for < 0.40; *fair* for 0.40 — 0.59; *good* for 0.60—0.74; and *excellent* for 0.75—1.00. Rather than reporting ICC agreement, which reflects both rank order agreement and intercept differences (e.g. also accounts for between-scanner discrepancies in absolute volumes), we also conducted a more detailed investigation in which we tested our ability to predict ‘unseen’ 1.5T values from 3T data using linear regression. To estimate the generalisation performance of a linear model, we computed the required slopes and intercepts for all imaging variables, and used 10-fold cross-validation to iteratively estimate a linear fit on 9/10^th^ of the data, applying prediction for the held-out fold and reporting average model fit (predicted R^2^). All imaging measures were modelled and estimated separately.

In order to assess if larger GM regions resulted in higher between-scanner consistency than smaller regions, we also reported the correlation between region volume (mean value of 1.5T and 3T volumes) and the regional ICCs. For the cortical surface vertex analyses (volume, surface area and thickness), in addition to providing regional maps of ICCs and percent difference, we performed linear regression between 1.5T and 3T values and computed uncorrected p-value maps to indicate areas of difference between scanners. To illustrate the impact of smoothing, we also provide average ICCs across the cortical mantle for volume, area and thickness smoothed with a 0, 5, 10, 15, 20, and 25 mm FWHM kernel.

## Results

Table 1 summarises the between-scanner statistics at each level of analysis. Broadly, we found a wide range (−13.2 to 39.1%) in the absolute difference between imaging measures at 1.5T and 3T. Figure 1 shows horizontal slices of two participants imaged at 1.5T alongside the equivalent slice at 3T. We observed different contrasts for skull, CSF, GM and WM between field strengths. Discrepancies in both the GM-WM boundary and the GM-CSF boundary were visible between field strengths and it was apparent that more GM and WM was visible at 3T than at 1.5T. Gibbs ringing artefacts were apparent at 1.5T but much less so at 3T.

**Table 1.** Summary of the between-scanner comparison performed at various levels of analysis using participants scanned at both 1.5T and 3.0T: Mean values, between-scanner differences and intraclass correlation coefficient (ICC). T1-weighted images were segmented using FreeSurfer 6.0. AD, RD and MD are measured in ×10^−3^ mm^2^ /s. AD=axial diffusivity; CSF=cerebrospinal fluid; FA=fractional anisotropy; GM=grey matter; MD=mean diffusivity; RD=radial diffusivity; WM=white matter.

|  |  | 1.5T mean | 3.0T mean | between-scanner difference (%) |  | ICC |
| --- | --- | --- | --- | --- | --- | --- |
| Global T1-weighted volumetric measures (cm<sup>3</sup>) | Supratentorial volume | 880.575 | 948.232 | 67.657 | (7.7) | 0.953 |
|  | Total CSF volume | 58.855 | 56.239 | -2.616 | (-4.4) | 0.993 |
|  | Total GM volume | 535.118 | 582.489 | 47.370 | (8.9) | 0.915 |
|  | Subcortical GM volume | 46.056 | 48.920 | 2.864 | (6.6) | 0.851 |
|  | Cortical GM volume | 392.928 | 432.806 | 39.878 | (10.2) | 0.892 |
|  | Cerebral WM volume | 386.890 | 413.609 | 26.719 | (7.0) | 0.824 |
|  |  | 1.5T mean | 3.0T mean | mean between-scanner difference (%) |  | mean ICC (SD) |
| Regional GM measures (16 subcortical, 68 cortical regions) | Subcortical volume (cm<sup>3</sup>) | 2.760 | 2.969 | 0.209 | (7.3) | 0.592 (0.174) |
|  | Cortical volume (cm<sup>3</sup>) | 5.793 | 6.395 | 0.602 | (10.8) | 0.750 (0.136) |
|  | Cortical surface area (cm<sup>2</sup>) | 22.047 | 25.081 | 3.034 | (12.3) | 0.733 (0.152) |
|  | Cortical thickness (mm) | 2.330 | 2.419 | 0.089 | (4.4) | 0.504 (0.134) |
|  |  | 1.5T mean | 3.0T mean | between-scanner difference (%) |  | ICC |
| Global dMRI measures in cerebral WM | AD | 1.041 | 1.103 | 0.062 | (6.0) | 0.776 |
|  | RD | 0.653 | 0.566 | -0.086 | (-13.2) | 0.882 |
|  | MD | 0.782 | 0.745 | -0.037 | (-4.7) | 0.867 |
|  | FA | 0.312 | 0.415 | 0.103 | (33.0) | 0.740 |
|  |  | 1.5T mean | 3.0T mean | mean between-scanner difference (%) |  | mean ICC (SD) |
| Regional dMRI measures in 12 WM tracts | MD | 0.793 | 0.740 | -0.054 | (-5.8) | 0.564 (0.193) |
|  | FA | 0.383 | 0.520 | 0.137 | (37.4) | 0.475 (0.127) |
|  | gMD | -0.102 | -0.064 | 0.044 | — | 0.850 — |
|  | gFA | 0.104 | 0.021 | -0.014 | — | 0.769 — |
|  |  | 1.5T mean | 3.0T mean | between-scanner difference (%) |  | ICC |
| Global network measures | Mean edge (MD) | 0.701 | 0.733 | 0.032 | (4.7) | 0.612 |
|  | Network efficiency (MD) | 0.492 | 0.493 | 0.001 | (0.3) | 0.888 |
|  | Network clustering coef (MD) | 0.526 | 0.498 | -0.028 | (-5.3) | 0.883 |
|  | Mean edge (FA) | 0.350 | 0.484 | 0.134 | (39.1) | 0.680 |
|  | Network efficiency (FA) | 0.248 | 0.328 | 0.080 | (32.6) | 0.794 |
|  | Network clustering coef (FA) | 0.248 | 0.318 | 0.070 | (28.6) | 0.799 |
|  |  | 1.5T mean | 3.0T mean | mean between-scanner difference (%) |  | mean ICC (SD) |
| Network connections (1071 connections) | MD-weighted | 0.701 | 0.733 | 0.032 | (4.6) | 0.280 (0.305) |
|  | FA-weighted | 0.350 | 0.484 | 0.134 | (38.4) | 0.275 (0.242) |

**Figure 1.**
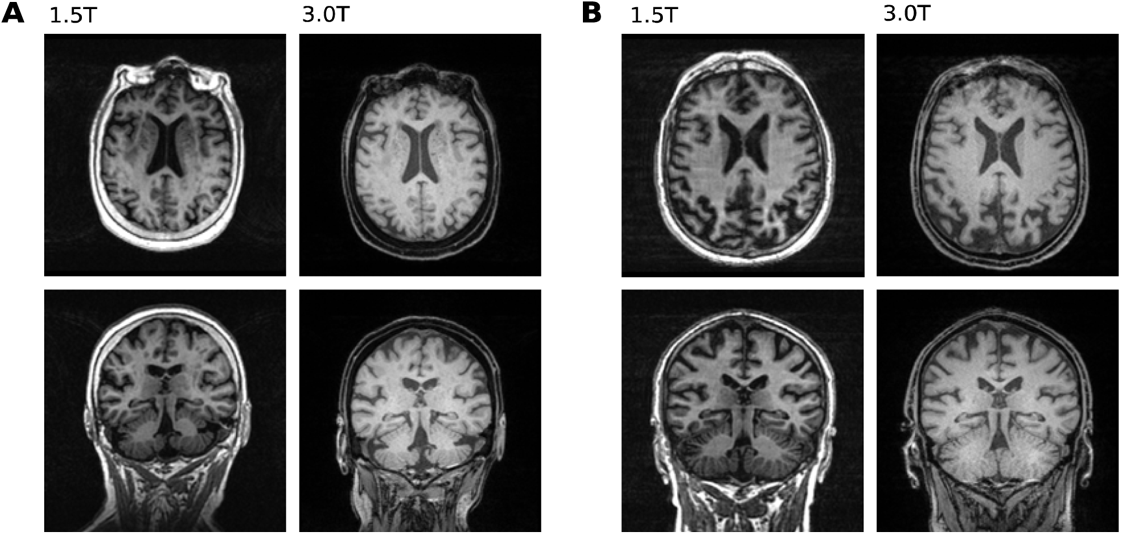
Axial and coronal T1-weighted slices at both 1.5T and 3T of one participant where the between-scanner supratentorial volume difference was measured at 55.86 cm^3^ (A) and another where supratentorial volume difference was 113.67 cm^3^ (B). The slices shown are in native T1 space (not co-registered) and were matched between scanners as closely as possible. Image intensity ranges were adjusted for visualisation.

### Between-scanner agreement of global volumetric measures

Supratentorial, GM and WM were estimated as 6.6% to 10.2% greater at 3T than at 1.5T. In particular, GM volumes were 8.9% greater for total GM, 10.2% greater for cortical GM and 6.6% greater for subcortical GM. WM volume was estimated as 7.0% greater at 3T than 1.5T. Conversely, total CSF volume was estimated as 4.4% lower at 3T than 1.5T. Scatter plots (Figure 2A) indicated that the between scanner relationships were largely linear (slopes between 0.688 and 1.048), and the Bland-Altman plots showed that there were few participants > 2 SD from the mean difference (Figure 2B).

**Figure 2.**
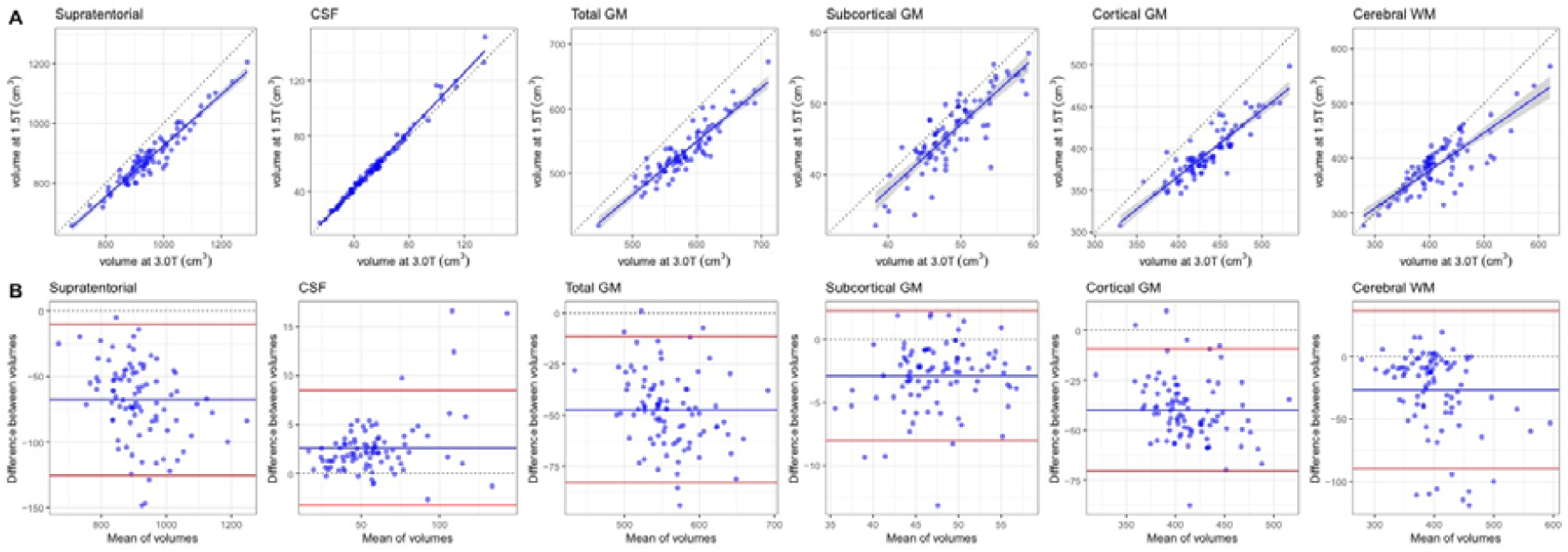
Between-scanner comparison of T1-weighted data: A) Scatter plots of six volumetric measures identified using FreeSurfer 6.0 for 91 participants scanned at both 1.5T and 3.0T, where the continuous blue line shows linear fit with 95% CI. B) Bland-Altman plots of the same six measures showing the mean of between-scanner volumes and the difference between these volumes where the blue line indicates the mean and the red lines represent ± 2 SDs. CSF=cerebrospinal fluid; GM=grey matter; WM=white matter.

Between-scanner consistency of global volumes, as assessed by ICC was considered excellent for all six global volumetric measures (ICCs ≥ 0.824; Table 1). Notably, total CSF volume had near perfect agreement between scanners (ICC = 0.993). Consistency was also excellent for supratentorial volume (ICC = 0.953), total GM (ICC = 0.915), cortical GM (ICC = 0.892), subcortical GM (ICC = 0.851), and WM volume (ICC = 0.824). Furthermore, consistency was good to excellent (ICC range: 0.680 to 0.993) for all global volumetric measures estimated by the FreeSurfer volumetric processing stream (Supplementary Table 1).

### Between-scanner agreement of grey matter measures

Mean values and between-scanner differences for cortical regions (volume, thickness and surface area) and subcortical volumes are reported in Supplementary Tables 2—5 and summarised in Table 1. The volumes of the 68 cortical regions were measured as 10.8% greater at 3T than at 1.5T on average (range: -7.0% to 28.6%). The volumes of the 16 subcortical regions were measured as 7.3% greater at 3T than at 1.5T (range: -21.0% to 27.6%). Cortical surface areas were measured as 12.3% larger at 3T than at 1.5T (range: -12.0% to 29.5%). Cortical thicknesses were measured as 4.4% (range: -4.6% to 12.3%) or 0.089 mm thicker on average at 3T than at 1.5T.

Figure 3 shows the between-scanner ICC consistency in volume, surface area and cortical thickness. Consistency was variable across measures and regions (ICC range: 0.142 to 0.930). For the 68 cortical regions the mean ICC for volume was 0.750 (range: 0.340 to 0.921) with consistency rated as excellent for 40 regions, good for 20 regions, fair for 6 regions and poor for the left/right frontal pole. Between-scanner consistency for cortical surface area was similar to cortical volume with a mean ICC of 0.733 (range: 0.335, 0.930). ICCs for surface area were rated as excellent for 40 regions, good for 14 regions, fair for 12 regions and poor for the left entorhinal and right insula. ICCs for cortical thickness were ∼0.2 lower than either cortical volume or surface area, with a mean ICC of 0.504 (range: 0.142 to 0.783) and consistency rated as excellent for only the right precuneus, good for 16 regions, fair for 40 regions and poor for 11 regions. Agreement was broadly similar between hemispheres for all regions (Figure 3). The ICC values and cortical properties were moderately correlated (volume *r* = 0.431, *p* < 0.001; area *r* = 0.275, *p* = 0.023; thickness *r* = 0.246, *p* = 0.043), which indicated that ICCs were generally lower for smaller regions, e.g. frontal pole and pallidum. Consistency for subcortical volumes were lower (mean ICC = 0.592, range: 0.217 to 0.853). ICCs for subcortical regions were rated as excellent for four regions, good for five regions, fair for 5 regions and poor for the left/right pallidum.

**Figure 3.**
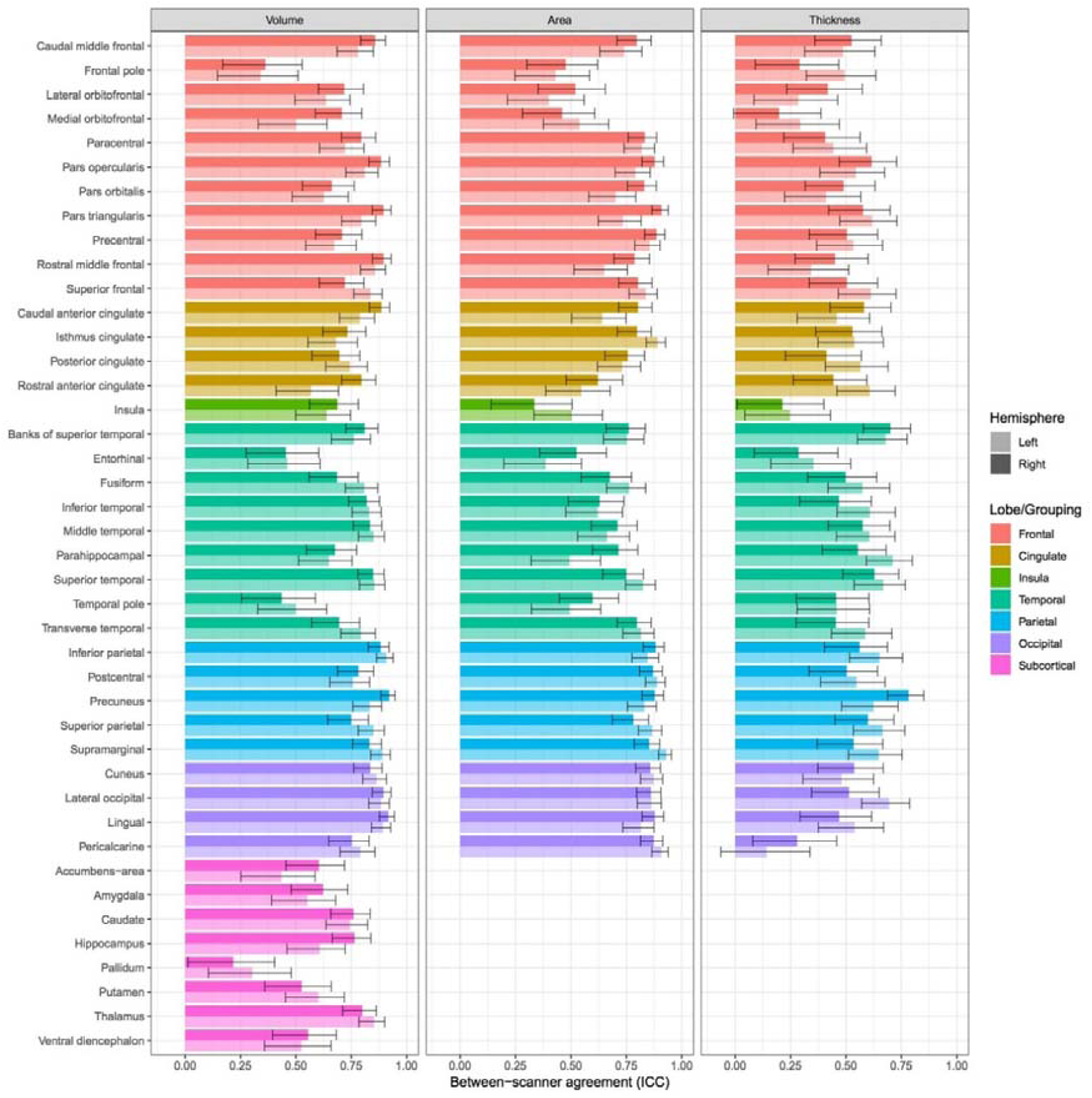
Consistency (ICC scores and estimated 95% CI) between 1.5T and 3.0T acquisitions for 84 grey matter regions identified by FreeSurfer 6.0 (N = 91) measuring volume, surface area and thickness. Surface area and thickness were not computed for subcortical regions.

### Between-scanner agreement in vertex-wise cortical measures

Figure 4 shows the between-scanner differences and ICCs for volume, area and thickness for data smoothed at 20 mm FWHM. Volume, surface area and thickness were all measured as greater at 3T than 1.5T. The mean between-scanner difference at vertex level was 0.132 mm^3^ (11.5% greater at 3T) for volume, 0.065 mm^2^ (14.1% greater at 3T) for surface area and 0.092 mm (4.8% thicker at 3T) for cortical thickness.

**Figure 4.**
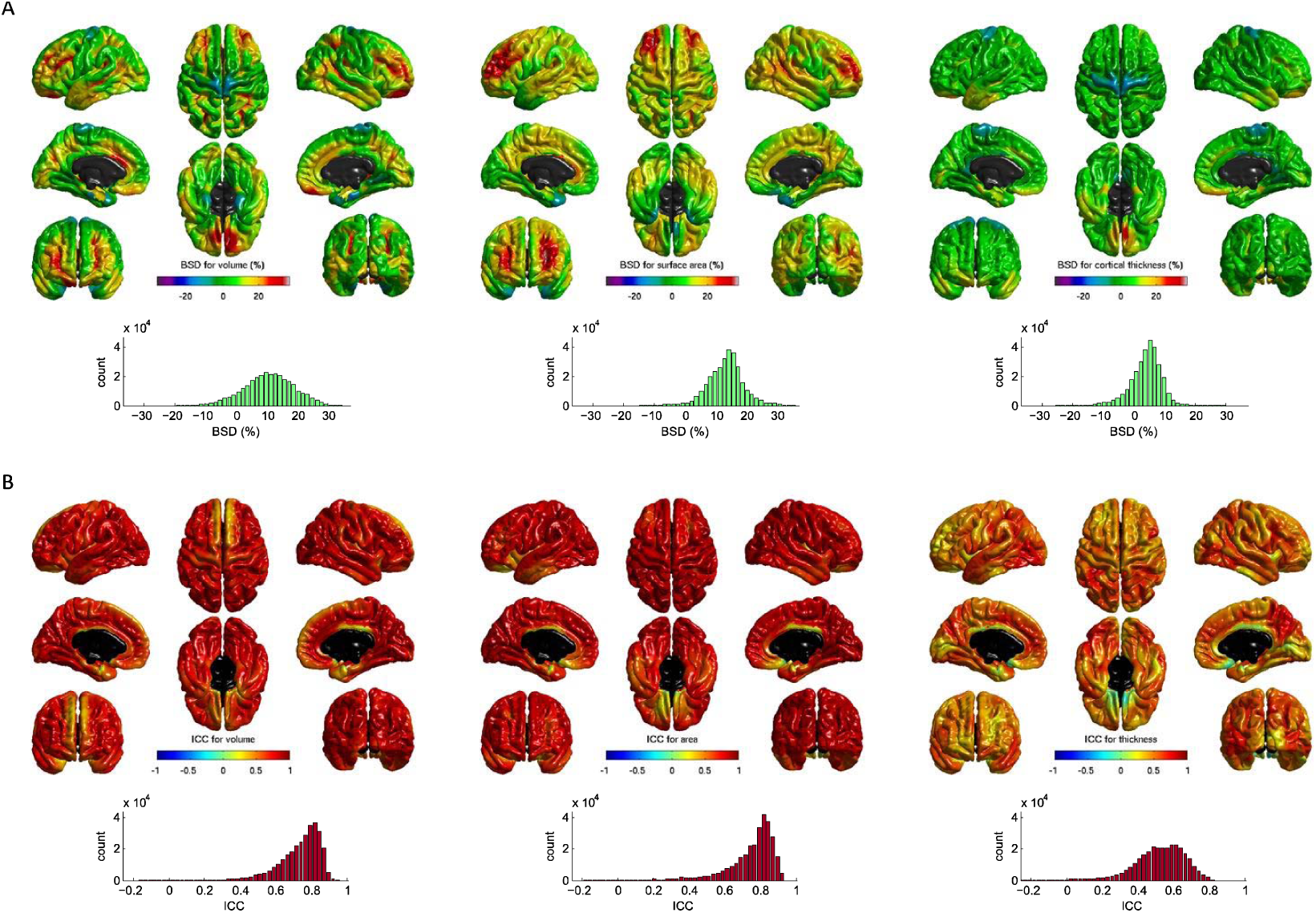
Between-scanner comparison of cortical volume, surface area and thickness for 91 participants imaged at both 1.5T and 3.0T: A) heatmaps show the between-scanner difference (BSD) expressed as percent change from 1.5T values at each cortical vertex location with corresponding histograms below; B) Intraclass correlation coefficient (ICC) of the same three measures with corresponding histograms. Processing was performed by FreeSurfer 6.0 and spatial smoothing using FWHM at 20 mm.

When computed across all vertices, the mean ICC values were broadly in line with those for the atlas-based regions reported above (Table 1): 0.747 (95% inter-percentile range [IPR]: 0.435 to 0.894) for cortical volume, 0.763 (95% IPR: 0.379 to 0.915) for surface area, and 0.535 (95% IPR: 0.192 to 0.761) for cortical thickness. Excellent between-scanner consistency (ICC > 0.75) was observed for 56.9% of individual vertices for volume and 64.6% of vertices for surface area, but this figure was substantially lower for cortical thickness (3.2% of vertices).

Scanner effects were somewhat regionally heterogeneous. Volumetric differences in the ICCs in the superior frontal lobe are mainly contributed to by lower ICCs for thickness, whereas lower volumetric ICCs in orbital frontal, cingulate and medial temporal regions were common to both area and thickness. Additionally, between-scanner contrasts for these three measures (Supplementary Figure 1), indicated that for our sample most cortical vertices were not significantly different between scanners (*p* < 0.05, uncorrected). Small areas of significant difference were observed in dorsal precentral gyrus (volume and thickness) and temporal poles (volume and area).

Between-scanner consistency increased as greater levels of FWHM smoothing were applied to the vertex level data (Supplementary Figure 2). For unsmoothed data, the mean ICCs of the three measures were between 0.150 and 0.403 but at the highest level of smoothing (25 mm) the mean ICCs were between 0.540 and 0.772. Smoothing had a larger effect on the mean ICCs for volume and surface area than for thickness.

### Between-scanner agreement of global dMRI measures

Figure 5 shows both scatter plots and corresponding Bland-Altman plots for four global dMRI measures (AD, RD, MD and FA) measured in cerebral WM. RD was measured as 13.2% lower and MD as 4.7% lower at 3T than 1.5T. AD was measured as 6.0% higher and FA as 33.0% higher at 3T than 1.5T. Scatter plots (Figure 5) indicated that the between scanner relationships were largely linear (slopes between 0.624 and 0.937). Additionally, Bland-Altman plots showed that there were very few participants > 2 SD difference from the mean difference.

**Figure 5.**
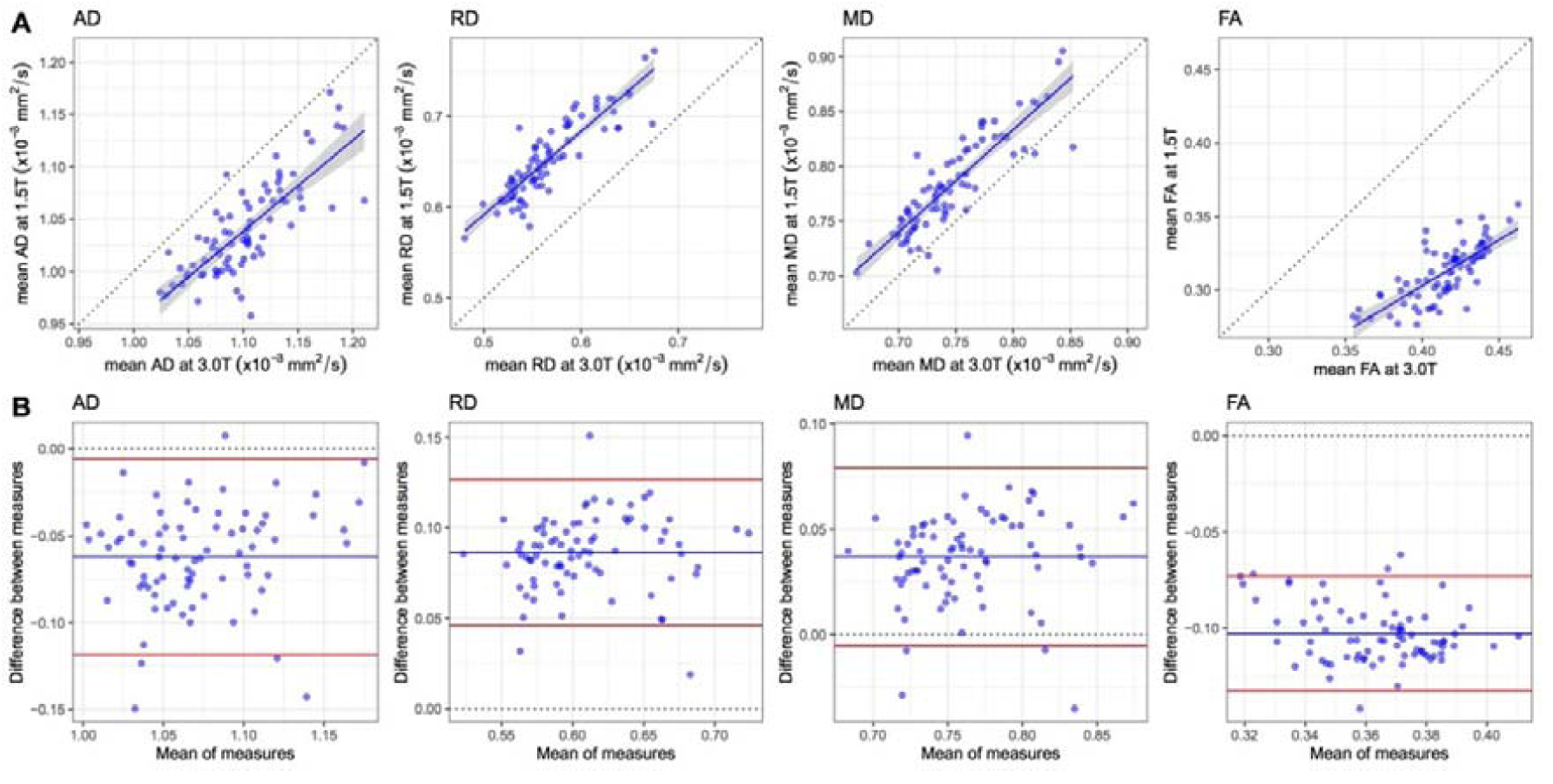
Between-scanner differences of four water diffusion measures, namely, axial diffusivity (AD), radial diffusivity (RD), mean diffusivity (MD) and fractional anisotropy (FA), measured in cerebral white matter for 79 participants scanned at both 1.5T and 3.0T: A) Scatter plots where the continuous blue line shows linear fit with 95% CI. B) Bland-Altman plots of the same four measures showing the mean of between-scanner volumes and the difference between these volumes where the blue line indicates the mean and the red lines represent ± 2 SDs.

Despite the differences in absolute levels, between-scanner consistency was considered excellent (RD ICC = 0.882; MD ICC = 0.867; AD ICC = 0.776), or good (FA ICC = 0.740; Table 1). In a supplementary analysis, we observed that ICCs were ∼0.1 lower in WM than when the same four measures were sampled across the whole-brain (Supplementary Table 6). The lower values for WM could be explained by the discrepancy in GM/WM segmentation between 1.5T and 3T, by which the whole-brain measures were unaffected.

### Between-scanner agreement in major white matter tracts

The mean values and the between-scanner differences of 12 white matter tracts are reported in Supplementary Table 7 and summarised in Table 1. Supplementary Figures 3 & 4 show scatter plots and Bland-Altman plots for these tracts. Visual inspection of the probability maps of each tract generated by probabilistic tractography revealed that streamlines more coherently followed the anatomical pathways at 3T than at 1.5T (Figure 6A – maps created using all data, prior to removal of QC fails), presumably due to the higher signal-to-noise and improved distortion correction at the higher field strength. Visual quality checking and exclusion of individual tracts identified more aberrant streamlines across subjects at 1.5T than at 3T with a tract success rate of 91.6—98.9% at 1.5T and 95.3—100% at 3T.

**Figure 6.**
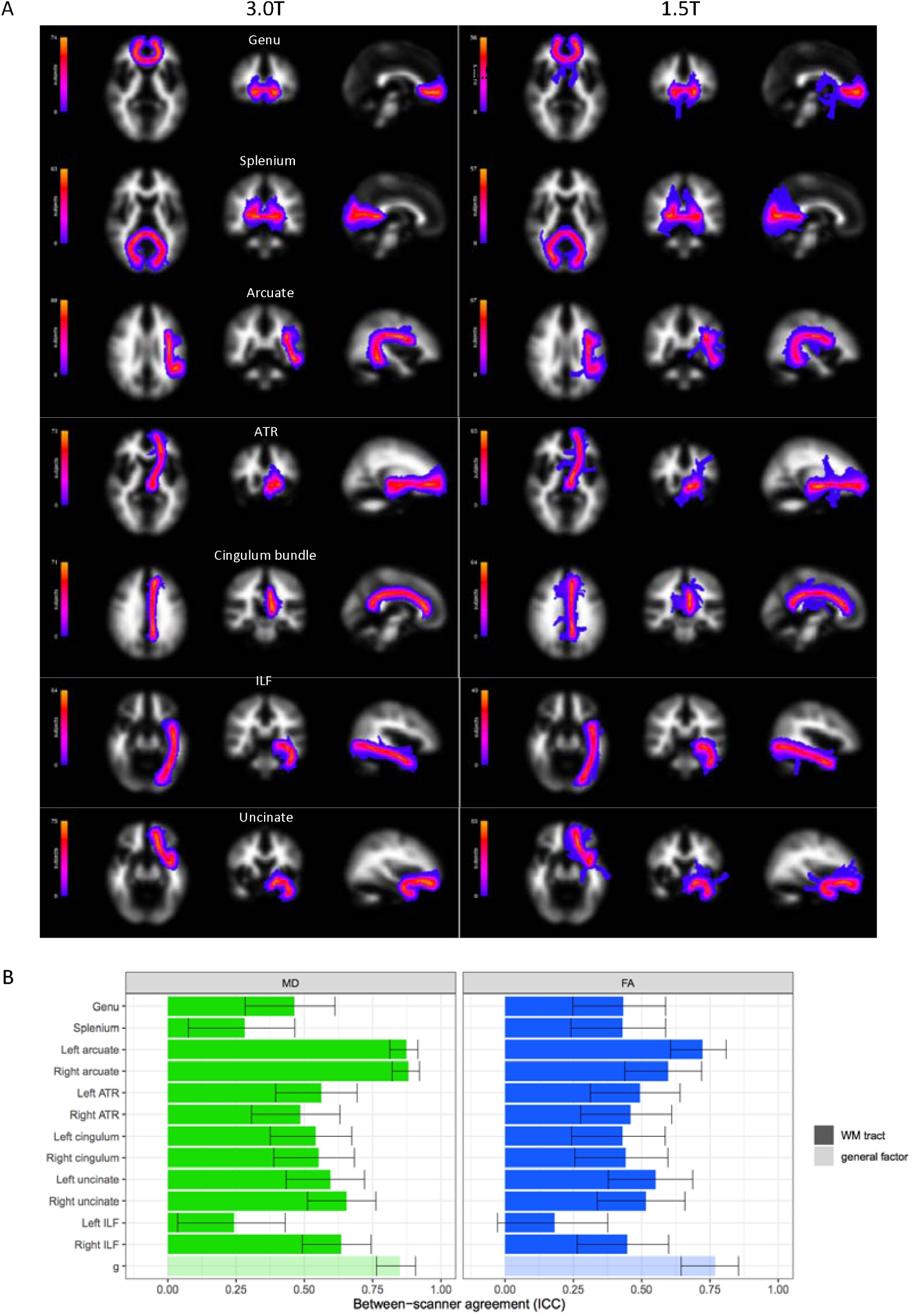
Between-scanner comparison of 12 white matter tracts in 90 participants: A) Anatomical probability maps for both 3.0T and 1.5T showing the streamline density of each tract (left-side only for bilateral tracts) across participants for whom validated tract data was available; B) Consistency (ICC and estimated 95% CI) between 1.5T and 3.0T acquisitions for 12 tracts (and their general factors extracted from the tract data) identified by probabilistic neighbourhood tractography and measuring both mean diffusivity (MD) and fractional anisotropy (FA). ATR=anterior thalamic radiations; ILF=inferior longitudinal fasciculus.

Across all tracts, FA was consistently higher at 3T (mean: 37.4%; range: 23.6 to 48.6%), and MD consistently lower (mean: 5.8%, range: -15.1 to 1.0%), with only the right cingulum bundle having a 1.0% increase in MD at the higher field strength. We also applied the 1.5T pipeline to 3T data from ten subjects and found that for the 12 tracts the FA values were measured as 3.4—24.8% higher using the 3T pipeline, suggesting that the apparent increase in FA at 3T was partly driven by the new FSL tools used in the 3T tractography pipeline.

Overall agreement was slightly better for MD (mean ICC of 0.564; range 0.243 to 0.881) than for FA (mean ICC of 0.475; range 0.182 to 0.723; Figure 6B). Between-scanner ICC consistency was rated as fair for the majority of tracts: 6/12 in MD and 8/12 tracts in FA. Consistency was excellent for only MD in the left and right arcuate (ICCs ≥ 0.873). Consistency was good for MD in the right uncinate and right ILF and for FA in the left arcuate. Consistency was poor (ICCs ≤ 0.282) for the left ILF (MD and FA) and for the splenium (MD).

General factors (gMD and gFA) of the 12 tracts were extracted using principal component analysis (loadings of the first unrotated principal component are listed in Supplementary Table 8). For gMD, the first unrotated principal component explained 44% of the variance at 1.5T and 56% at 3T. For gFA, the first principal component explained 35% of the variance at 1.5T and 31% at 3T. Both gMD and gFA provided excellent between-scanner consistency (ICCs of 0.850 and 0.769, respectively), which was ∼0.3 greater than the mean ICC of the 12 tracts (Figure 6B & Supplementary Table 7).

### Between-scanner comparison of connectome

MD- and FA-weighted whole-brain networks were computed allowing 3570 possible WM connections for unthresholded networks but with only 1071 connections retained after consistency-thresholding at 30%. Between-scanner results for individual connection weights (edges) and three global graph-theoretic measures (mean edge weight, global network efficiency and network clustering coefficient) are shown in Supplementary Table 9 and summarised in Table 1.

For unthresholded networks, the connection density was 79.6% greater at 3T than 1.5T (mean network sparsity: 0.599 [SD = 0.037] for 1.5T; 0.334 [SD = 0.048] for 3T), meaning that considerably more interregional WM connections were identified at the higher field strength, presumably due to higher signal-to-noise. However, after network thresholding, which retained only the top 30% most consistent connections across subjects, each participant’s network was constrained to have a sparsity of ∼0.7. Separate thresholds were applied at 1.5T and 3T which resulted in a different set of connections after thresholding. However, we found that there was a high overlap in the connections retained (ICC = 0.687 or 835/1307 matching connections) when comparing the binary masks obtained from the thresholding procedure between the two field strengths.

For MD weighted networks, mean edge weight was measured as 4.7% greater, network efficiency as 0.3% greater and network clustering coefficient as 5.3% lower at 3T than 1.5T. For FA weighted networks, mean edge weight was 39.1% greater, network efficiency was 32.6% greater and network clustering coefficient was 28.6% greater at 3T than 1.5T. Figure 7A&B shows the between-scanner results for these network metrics for both MD and FA networks. Consistency was rated as good to excellent for all global metrics (ICC range: 0.612 to 0.888). The greatest consistency was for network efficiency with MD (ICC = 0.888) and network clustering coefficient with MD (ICC = 0.883). FA-weighted metrics were rated as excellent for network clustering coefficient (ICC = 0.799) and network efficiency (ICC = 0.794). The lowest consistency for these measures, which was rated as good, was for mean edge weight with FA (ICC = 0.612) and MD (ICC = 0.680). Despite these differences in between-scanner consistency for the three metrics, we note high collinearity among the three graph-theoretic measures (*r* > 0.796 for MD; and *r* > 0.943 for FA).

**Figure 7:**
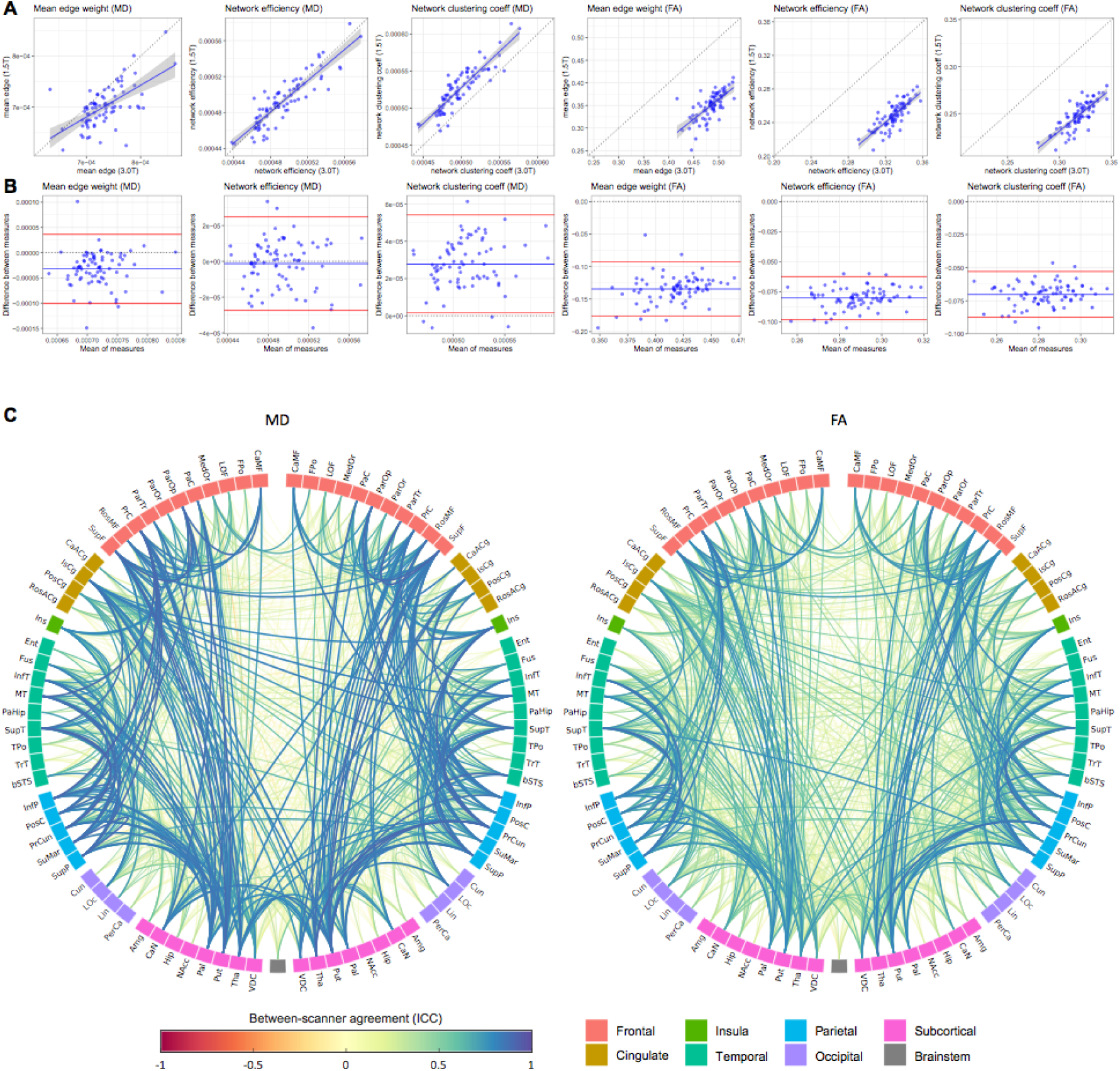
Between-scanner results for whole-brain structural networks using 85 nodes with connection strength weighted by both MD and FA and computed using 79 participants scanned at both 1.5 and 3.0T. Networks were thresholded to retain the top 30% consistent connections across participants. A) Scatter plots for three global network metrics for both MD and FA networks, where the continuous blue line shows linear fit with 95% CI. B) Bland-Altman plots of the same network metrics showing the mean of between-scanner volumes and the difference between these volumes where the blue line indicates the mean and the red lines represent ± 2 SDs; C) Anatomical network plots for FA- and MD-weighted networks, where link colour and thickness represents the ICC for each connection (edge). Node abbreviations are listed in Supplementary Table 10.

The ICCs for each of the 1071 individual connections which were retained following 30% network thresholding are shown in Figure 7C. Overall between-scanner consistency was poor for both MD and FA networks (mean ICCs ≤ 0.280; Table 1). For FA, the mean ICC was 0.275 although the 95% IPR was broad (−0.051 to 0.795). This corresponded to a proportion of excellent/good/fair/poor of 5.4/9.0/12.4/73.2%. For MD, the mean ICC was 0.280 (95% IPR: -0.095 to 0.870) corresponding to a proportion of excellent/good/fair/poor of 14.5/8.0/6.5/71.1%. Whereas 30% thresholded networks achieved better between-scanner consistency (mean ICCs ≤ 0.280) than unthresholed networks (mean ICCs ≤ 0.142), this result was driven by the large difference in network sparsity between scanners, i.e. there are many more zero-valued connections (marking an absence of connection between regions) at 1.5T compared to 3T. When all zero-valued connections (Supplementary Figure 5) were excluded and ICCs were computed for the 428 network connections (12% of all possible connections), which had a nonzero value in every participant, then the between-scanner consistency was considerably higher (mean ICC of 0.647 for MD and 0.533 for FA).

### Prediction of ‘unseen’ 1.5T imaging variables from 3T data

Slopes and intercepts from a linear fit of global and regional imaging variables between scanners are reported in Supplementary Tables 1-7 & 9, alongside the predicted model fit (R^2^) obtained from 10-fold cross-validation with a linear model.

The range of predicted model fit was variable across all imaging measures (0.155—0.991) but the highest R^2^ values indicted that differences between scanners could be virtually eliminated (almost perfect prediction) for global volumetric measures and large brain structures. For global T1 volumetric measures (Supplementary Table 1), the R^2^ range was 0.615—0.991 with estimated intracranial volume having the lowest and CSF having the highest R^2^. For volumetric variables derived from FreeSurfer volumetric and subcortical processing (Supplementary Table 2), the R^2^ range was 0.130— 0.990 with left pallidum lowest and right lateral ventricle highest. For the cortical measures (Supplementary Tables 3—5), the R^2^ range was 0.202—0.860 for volume, 0.195—0.852 for surface area and 0.155—0.684 for cortical thickness with the right frontal pole, insula and entorhinal areas scoring lowest and right precuneus obtaining the highest R^2^ for volume and area. For global dMRI measures (Supplementary Table 6) the R^2^ range was 0.648—0.863 with FA in WM lowest, and MD in whole-brain highest. For WM tracts (Supplementary Table 7) the R^2^ range was 0.196—0.845 with left ILF (FA) lowest and left arcuate (MD) highest. For global network metrics (Supplementary Table 9) the R^2^ range was 0.483—0.827, with mean edge weight (MD) lowest and network clustering coefficient (MD) having the highest R^2^.

## Discussion

In one of the largest between-scanner comparisons to date, we report previously lacking information on a wide range of structural brain measures in an exclusively older group of participants. We found excellent levels of consistency (ICC > ∼0.75) between the 1.5T and 3T scanners for the largest brain structures (whole-brain, ventricular and tissue volumes; global dMRI measures in WM; and global network metrics) that were similar to same-scanner test-retest studies (Buchanan et al., 2014; Iscan et al., 2015; Luque Laguna et al., 2020; Melzer et al., 2020). We noted that there were overall mean shifts in the absolute levels of most measures between 1.5T and 3T: volumetric measures and thickness appeared larger at 3T, RD and MD were lower, and AD and FA were higher at 3T, consistent with prior observations from smaller studies on single metrics (Chu et al., 2017; Han et al., 2006; Heinen et al., 2016; Pfefferbaum et al., 2012), but not others (West et al., 2013). Regression-based correction for scanner (using intercept differences) effectively eliminated scanner differences in unseen (hold-out) data for global brain measures, giving similar (and sometimes higher) agreement than might be expected from same-scanner test-retest data: global measures could be accurately predicted in line with 1.5T values from 3T data using 10-fold cross-validation.

Interestingly, both grey and whiter matter tissue volumes appeared larger at 3T than at 1.5T, but CSF volume was slightly smaller. Contributing factors are likely to include a combination of higher tissue contrast (resulting in differences in the tissue-CSF boundary), different scanner-specific geometric distortions and a slight difference in T1-weighted voxel dimensions. More numerous sampling instances along a complex surface may result in both superior estimation (c.f. Cavalieri), and the ‘coastline paradox’, whereby complex shapes appear larger when measured with greater fidelity (c.f. Richardson; (Napolitano, Ungania, & Cannat, 2012). This clearly has important implications for cross-scanner analyses that use ICV or CSF correction to measure atrophic change in global tissue volumes from cross-sectional data with different voxel dimensions – lower field strengths may potentially result in higher estimates of atrophy.

As would be expected, between-scanner agreement decreased as the granularity increased from large brain structures to include smaller regional imaging variables. Scanner agreement at the regional level was similar or slightly lower than prior same-scanner work, such as for cortical regional measures (Boekel et al., 2017; Clayden et al., 2009; Liem et al., 2015; Luque Laguna et al., 2020; Madan & Kensinger, 2017; Srinivasan et al., 2020). We also found that smaller GM regions typically had poorer between-scanner agreement than large regions; this between-scanner finding corresponds well with the known relationship between reliability and region size observed in same-scanner work (Iscan et al., 2015; Tustison et al., 2014). This finding indicates that in this specific case, scanner differences may not contribute a substantial amount of additional noise to the noise reliability typically seen in test-retest settings. It also contributes more generally to the literature on the merits and drawbacks of increasing cortical atlas granularity for the reliability of the structural connectome (de Reus & van den Heuvel, 2013) or structural-functional correspondence (Messé, 2020). Additionally, a recent 1.5-3T field strength comparison (N = 113), reported a broadly similar pattern for regional reliability of FreeSurfer segmentation (Srinivasan et al., 2020). The authors of that study also identified a bias in the FreeSurfer procedure for under segmentation of subcortical structures, particularly hippocampal volumes, in older subjects.

Our vertex-wise cortical analyses were valuable in that they show that ICCs increase with greater smoothing and show a pattern of between-scanner ICC consistency which is agnostic to boundaries imposed by a particular cortical atlas. Prior findings suggest that cortical thickness generally shows lower reliability than either volume or surface area in a same-scanner setting (Iscan et al., 2015), with which our findings are consistent. Interestingly, although the percent differences between 1.5T and 3T data were wider for volume and surface area (especially prevalent in lateral and orbital frontal, cingulate and posterior temporal areas) than for thickness, ICCs were very much lower for thickness than either volume or area. Thus, whereas the overall volume or area of cortex identified is proportionally higher at 3T than for thickness, this overestimation is far more systematic (the rank order is better preserved across scanners) than for thickness. It is possible that thickness appears less reliable between 1.5-3T because the two dimensions upon which it relies (grey-white and grey-CSF boundaries) to derive sub-millimetre measurements are those that would be affected by contrast differences between field strengths.

With respect to dMRI data, the increase in FA and AD, and decrease in RD and MD between 1.5T and 3T, as well as the higher number of WM inter-regional connections may also be indicative of superior signal-to-noise (a better fit of the diffusion tensor). However, these differences may be also partly explained by improved distortion correction at 3T. On this latter factor, the application of a modified pipeline for the PNT-identified WM tracts (Tractor v2.6 with FSL v4 at 1.5T and Tractor v3.3 with FSL v5) was necessary to work with multi-shell data for which the prior versions were not optimised, and to apply the more advanced tools in eddy-current distortion and susceptibility corrections that we would be using in future study waves at 3T. This is likely to have provided an additional source of inconsistency for the PNT-identified WM tract analyses, which were generally poorer than ICCs from similar methods in same-scanner designs which report ICCs > 0.54 (Boekel et al., 2017; Clayden et al., 2009; Luque Laguna et al., 2020). Indeed, the contribution of pipeline differences was borne out in our supplementary analyses: applying a different pipeline to 3T substantially affected dMRI measures in a small sample of our participants and FA increased substantially in WM tracts using more recent processing algorithms, which provided better distortion corrections (FA tends to increase with better distortion correction; Yamada et al., 2014). Nevertheless, our findings in the main analyses still indicated ‘fair’ consistency, with poorer agreement found for smaller tracts which involved fewer streamlines and were generally found close to the ventricles; these were more likely to suffer from partial CSF contamination in some streamlines.

Global network metrics derived from the structural connectome showed good to excellent consistency, comparable to same-scanner results (Buchanan et al., 2014; Cheng et al., 2012). We found between-scanner consistency to be poor at the level of individual connections, though this was vastly improved when we accounted for differences in network sparsity (many more connections were identified at 3T than at 1.5T). Poor between-scanner consistency was not unexpected because the variability in T1-weighted regional segmentation and tractography both contribute to the variability in the resulting networks. Our results suggest that multi-scanner network analyses require careful consideration in the treatment of acquisition-specific network sparsities, such as the use of stringent thresholding or other de-noising methods (de Reus & van den Heuvel, 2013; Roberts et al., 2017).

### Limitations

The present study has several limitations which should be taken into account. Our aim was to determine between scanner differences in a sample of exclusively older subjects including those with representative age-related pathology. However, same-scanner test-retest variability has been shown to be greater in older subjects than in younger (Jovicich et al., 2009). Additionally, there was a relatively large interval (mean of 72 days) between scans, but even in older age it is unlikely that age-related structural changes can be reliably detected by MRI over such a short period (Resnick et al., 2000). In addition, direct comparison to prior between-scanner and same-scanner test-retest studies is problematic because different statistics are commonly used, including different formulations of the ICC.

The specific scanner configurations used in this work may limit the generalisability of the current findings given that between-scanner agreement can be influenced by scanner manufacturers, acquisition parameters and image processing software (Heinen et al., 2016; Jovicich et al., 2009; Tardif, Collins, & Pike, 2010; Wardlaw et al., 2012). Our study represents a change in field strength, manufacturer, acquisition and some necessary processing steps (e.g. for dMRI processing), such that we must be clear that the differences between scans cannot be only attributed to field strength. A previous between-scanner comparison showed that reliability is typically better when the same scanner manufacture was used than when different scanner manufactures were used (Jovicich et al., 2009). Some between-scanner volumetric variability must be attributed to the slight mismatch in T1-weighted voxel dimensions (only 3T voxels were isotropic). The spatial resolution used in our primary study was due to constraints on the scanning time for the required modalities. However, the FreeSurfer morphometric procedure was designed to be sequence-independent and involves interpolating T1 volumes to isotropic voxels before segmentation (Fischl, Salat, et al., 2004). Additionally, for longitudinal settings the more recent FreeSurfer longitudinal processing pipeline has been shown to obtain better cross-session reliability than the cross-sectional pipeline (Jovicich et al., 2013). Additionally, we did use openly-accessible and commonly-used methods across a large range of structural neuroimaging measures. Different versions of dMRI processing software were used as we needed to keep 1.5T acquisition and processing consistent with prior waves of our longitudinal study. We cannot use the newer FSL software tools in our 1.5T data as we do not acquire the necessary reverse phase-encoded volumes (https://fsl.fmrib.ox.ac.uk/fsl/fslwiki/topup).

We employed a straightforward linear regression approach using k-fold cross-validation, and therefore cannot rule out that promising scanner harmonization and calibration methods will not further improve cross-scanner reliability (Cetin Karayumak et al., 2019; Keshavan et al., 2016; Pinto et al., 2020; Tax et al., 2019). Finally, we judged that providing uncorrected, rather than corrected, p-values in the statistical test of scanner differences at the vertex level was more sensitive for the purpose of illustrating any potential differences. Nevertheless, there is relatively low statistical power here (e.g. Schönbrodt and Perugini, 2013) — even though this study represents one of the largest cross-scanner studies – potentially resulting in an underestimation of cross-scanner differences that may be apparent in larger meta- and mega-analytic settings.

## Conclusions

Longstanding longitudinal studies are torn between maintaining consistency of the MRI protocol and embracing improvements in scanner technology. The present study reports previously lacking cross-scanner results on a broad range of structural brain measures in a comparatively large sample of older participants. Global measures showed consistently good or excellent agreement, with lower agreement seen with increasing granularity of measurement, though in most cases these were still comparable to prior within-scanner test-retest results. Differences in the absolute level were prevalent, but we showed that, particularly for global measures, between-scanner variability could be effectively eliminated in unseen (hold-out) data using a k-fold cross-validation linear model. We conclude that low granularity measures of brain structure can be reliably measured between the different scanner manufacturers and field strengths tested. However, we recommend caution in combining high granularity information from different scanners. These data have useful implications for multi-centre meta- and mega-analyses combining data across hardware, software and field strengths (Van Den Heuvel et al., 2019), and provide much-needed information in an exclusively older age group which is underrepresented in this literature.

## Data Availability

Participant data can be accessed to researchers through a data request, and under a formal data sharing agreement, as outlined on the study website: https://www.ed.ac.uk/lothian-birth-cohorts/data-access-collaboration

## Acknowledgements

We thank the Lothian Birth Cohort 1936 members who took part in this study, and Lothian Birth Cohort 1936 research team members and radiographers who collected and checked data used in this manuscript. The LBC1936 and this research are supported by Age UK (Disconnected Mind project) and by the UK Medical Research Council [MRC; G0701120, G1001245, MR/M013111/1, MR/R024065/1]. CRB, SRC, MEB, IJD and EMT-D were also supported by a National Institutes of Health (NIH) research grant R01AG054628. JMW and IJD are also supported by a Wellcome Trust Strategic Award (Ref 104036/Z/14/Z). MCVH is funded by the Row Fogo Charitable Trust (grant No. BROD.FID3668413). EMT-D is a member of the Population Research Center at the University of Texas at Austin, which is supported by NIH center grant P2CHD042849. TCR is a member of the Alzheimer Scotland Dementia Research Centre supported by Alzheimer Scotland.

## References

Andersson, J. L. R., Skare, S., & Ashburner, J. (2003). How to correct susceptibility distortions in spinecho echo-planar images: Application to diffusion tensor imaging. NeuroImage. https://doi.org/10.1016/S1053-8119(03)00336-7

Andersson, J. L. R., & Sotiropoulos, S. N. (2016). An integrated approach to correction for off-resonance effects and subject movement in diffusion MR imaging. NeuroImage. https://doi.org/10.1016/j.neuroimage.2015.10.019

Behrens, T. E. J., Berg, H. J., Jbabdi, S., Rushworth, M. F. S., & Woolrich, M. W. (2007). Probabilistic diffusion tractography with multiple fibre orientations: What can we gain? NeuroImage, 34(1), 144–155. Retrieved from http://www.ncbi.nlm.nih.gov/pubmed/17070705

Behrens, T. E. J., Woolrich, M. W., Jenkinson, M., Johansen-Berg, H., Nunes, R. G., Clare, S., … Smith, S. M. (2003). Characterization and propagation of uncertainty in diffusion-weighted MR imaging. Magnetic Resonance in Medicine, 50(5), 1077–1088. Retrieved from http://www.ncbi.nlm.nih.gov/pubmed/14587019

Boekel, W., Forstmann, B. U., & Keuken, M. C. (2017). A test-retest reliability analysis of diffusion measures of white matter tracts relevant for cognitive control. In Psychophysiology. https://doi.org/10.1111/psyp.12769

Buchanan, C. R., Bastin, M. E., Ritchie, S. J., Liewald, D. C., Madole, J. W., Tucker-Drob, E. M., … Cox, S. R. (2020). The effect of network thresholding and weighting on structural brain networks in the UK Biobank. NeuroImage. https://doi.org/10.1016/j.neuroimage.2019.116443

Buchanan, C. R., Pernet, C. R., Gorgolewski, K. J., Storkey, A. J., & Bastin, M. E. (2014). Test-retest reliability of structural brain networks from diffusion MRI. NeuroImage, 86, 231–243. https://doi.org/10.1016/j.neuroimage.2013.09.054

Cetin Karayumak, S., Bouix, S., Ning, L., James, A., Crow, T., Shenton, M., … Rathi, Y. (2019). Retrospective harmonization of multi-site diffusion MRI data acquired with different acquisition parameters. NeuroImage. https://doi.org/10.1016/j.neuroimage.2018.08.073

Cheng, H., Wang, Y., Sheng, J., Kronenberger, W. G., Mathews, V. P., Hummer, T. A., & Saykin, A. J. (2012). Characteristics and variability of structural networks derived from diffusion tensor imaging. NeuroImage, 61, 1153–1164. https://doi.org/10.1016/j.neuroimage.2012.03.036

Chu, R., Hurwitz, S., Tauhid, S., & Bakshi, R. (2017). Automated segmentation of cerebral deep gray matter from MRI scans: Effect of field strength on sensitivity and reliability. BMC Neurology, https://doi.org/10.1186/s12883-017-0949-4

Cicchetti, D. V. (1994). Guidelines, Criteria, and Rules of Thumb for Evaluating Normed and Standardized Assessment Instruments in Psychology. Psychological Assessment, https://doi.org/10.1037/1040-3590.6.4.284

Clayden, J. D., King, M. D., Maniega, S. M., Bastin, M. E., Storkey, A. J., & Clark, C. A. (2011). TractoR: Magnetic resonance imaging and tractography with R. Journal of Statistical Software, https://doi.org/10.18637/jss.v044.i08

Clayden, J. D., Storkey, A. J., Maniega, S. M., & Bastin, M. E. (2009). Reproducibility of tract segmentation between sessions using an unsupervised modelling-based approach. NeuroImage. https://doi.org/10.1016/j.neuroimage.2008.12.010

de Reus, M. A., & van den Heuvel, M. P. (2013). Estimating false positives and negatives in brain networks. NeuroImage, 70, 402–409. https://doi.org/10.1016/j.neuroimage.2012.12.066

Deary, I J, Gow, A. J., Taylor, M. D., Corley, J., Brett, C., Wilson, V., … others. (2007). The Lothian Birth Cohort 1936: a study to examine influences on cognitive ageing from age 11 to age 70 and beyond. BMC Geriatrics, 7(1), 28.

Deary, Ian J., Gow, A. J., Pattie, A., & Starr, J. M. (2012). Cohort profile: The lothian birth cohorts of 1921 and 1936. International Journal of Epidemiology, https://doi.org/10.1093/ije/dyr197

Desikan, R. S., Ségonne, F., Fischl, B., Quinn, B. T., Dickerson, B. C., Blacker, D., … Killiany, R. J. (2006). An automated labeling system for subdividing the human cerebral cortex on MRI scans into gyral based regions of interest. NeuroImage, 31(3), 968–980. Retrieved from http://www.ncbi.nlm.nih.gov/pubmed/16530430

Fischl, B., Salat, D. H., Busa, E., Albert, M., Dieterich, M., Haselgrove, C., … Dale, A. M. (2002). Whole brain segmentation: Automated labeling of neuroanatomical structures in the human brain. Neuron, 33(3), 341–355. https://doi.org/10.1016/S0896-6273(02)00569-X

Fischl, B., Salat, D. H., Van Der Kouwe, A. J. W., Makris, N., Ségonne, F., Quinn, B. T., & Dale, A. M. (2004). Sequence-independent segmentation of magnetic resonance images. NeuroImage, 23 Suppl 1, S69–84. https://doi.org/10.1016/j.neuroimage.2004.07.016

Fischl, B., Van Der Kouwe, A., Destrieux, C., Halgren, E., Ségonne, F., Salat, D. H., … Dale, A. M. (2004). Automatically parcellating the human cerebral cortex. Cerebral Cortex, 14(1), 11–22. Retrieved from http://www.cercor.oupjournals.org/cgi/doi/10.1093/cercor/bhg087

Fushimi, Y., Miki, Y., Urayama, S. I., Okada, T., Mori, N., Hanakawa, T., … Togashi, K. (2007). Gray matter-white matter contrast on spin-echo T1-weighted images at 3 T and 1.5 T: A quantitative comparison study. European Radiology, https://doi.org/10.1007/s00330-007-0688-9

Grech-Sollars, M., Hales, P. W., Miyazaki, K., Raschke, F., Rodriguez, D., Wilson, M., … Clark, C. A. (2015). Multi-centre reproducibility of diffusion MRI parameters for clinical sequences in the brain. NMR in Biomedicine, https://doi.org/10.1002/nbm.3269

Gronenschild, E. H. B. M., Habets, P., Jacobs, H. I. L., Mengelers, R., Rozendaal, N., Van Os, J., & Marcelis, M. (2012). The Effects of FreeSurfer Version, Workstation Type, and Macintosh Operating System Version on Anatomical Volume and Cortical Thickness Measurements. PLoS ONE, 7(6), e38234. https://doi.org/10.1371/journal.pone.0038234

Gunter, J. L., Bernstein, M. A., Borowski, B. J., Ward, C. P., Britson, P. J., Felmlee, J. P., … Jack, C. R. (2009). Measurement of MRI scanner performance with the ADNI phantom. Medical Physics, https://doi.org/10.1118/1.3116776

Han, X., Jovicich, J., Salat, D., Van Der Kouwe, A., Quinn, B., Czanner, S., … Fischl, B. (2006). Reliability of MRI-derived measurements of human cerebral cortical thickness: the effects of field strength, scanner upgrade and manufacturer. NeuroImage, 32(1), 180–194. Retrieved from http://www.ncbi.nlm.nih.gov/pubmed/16651008

Heinen, R., Bouvy, W. H., Mendrik, A. M., Viergever, M. A., Biessels, G. J., & De Bresser, J. (2016). Robustness of automated methods for brain volume measurements across different MRI field strengths. PLoS ONE, https://doi.org/10.1371/journal.pone.0165719

Iscan, Z., Jin, T. B., Kendrick, A., Szeglin, B., Lu, H., Trivedi, M., … Delorenzo, C. (2015). Test-retest reliability of freesurfer measurements within and between sites: Effects of visual approval process. Human Brain Mapping, https://doi.org/10.1002/hbm.22856

Jenkinson, M., & Smith, S. (2001). A global optimisation method for robust affine registration of brain images. Medical Image Analysis, https://doi.org/10.1016/S1361-8415(01)00036-6

Jovicich, J., Czanner, S., Han, X., Salat, D., van der Kouwe, A., Quinn, B., … Fischl, B. (2009). MRI-derived measurements of human subcortical, ventricular and intracranial brain volumes: Reliability effects of scan sessions, acquisition sequences, data analyses, scanner upgrade, scanner vendors and field strengths. NeuroImage. https://doi.org/10.1016/j.neuroimage.2009.02.010

Jovicich, J., Marizzoni, M., Sala-Llonch, R., Bosch, B., Bartrés-Faz, D., Arnold, J., … Frisoni, G. B. (2013). Brain morphometry reproducibility in multi-center 3T MRI studies: A comparison of cross-sectional and longitudinal segmentations. NeuroImage. https://doi.org/10.1016/j.neuroimage.2013.05.007

Keshavan, A., Paul, F., Beyer, M. K., Zhu, A. H., Papinutto, N., Shinohara, R. T., … International Multiple Sclerosis Genetics Consortium. (2016). Power estimation for non-standardized multisite studies. NeuroImage. https://doi.org/10.1016/j.neuroimage.2016.03.051

Kruggel, F., Turner, J., & Muftuler, L. T. (2010). Impact of scanner hardware and imaging protocol on image quality and compartment volume precision in the ADNI cohort. NeuroImage. https://doi.org/10.1016/j.neuroimage.2009.11.006

Liem, F., Mérillat, S., Bezzola, L., Hirsiger, S., Philipp, M., Madhyastha, T., & Jäncke, L. (2015). Reliability and statistical power analysis of cortical and subcortical FreeSurfer metrics in a large sample of healthy elderly. NeuroImage. https://doi.org/10.1016/j.neuroimage.2014.12.035

Luque Laguna, P. A., Combes, A. J. E., Streffer, J., Einstein, S., Timmers, M., Williams, S. C. R., & Dell’Acqua, F. (2020). Reproducibility, reliability and variability of FA and MD in the older healthy population: A test-retest multiparametric analysis. NeuroImage: Clinical. https://doi.org/10.1016/j.nicl.2020.102168

Madan, C. R., & Kensinger, E. A. (2017). Test–retest reliability of brain morphology estimates. Brain Informatics, https://doi.org/10.1007/s40708-016-0060-4

Melzer, T. R., Keenan, R. J., Leeper, G. J., Kingston-Smith, S., Felton, S. A., Green, S. K., … Myall, D. J. (2020). Test-retest reliability and sample size estimates after MRI scanner relocation. NeuroImage. https://doi.org/10.1016/j.neuroimage.2020.116608

Messé, A. (2020). Parcellation influence on the connectivity-based structure–function relationship in the human brain. Human Brain Mapping, https://doi.org/10.1002/hbm.24866

Morey, R. A., Selgrade, E. S., Wagner, H. R., Huettel, S. A., Wang, L., & McCarthy, G. (2010). Scan-rescan reliability of subcortical brain volumes derived from automated segmentation. Human Brain Mapping, 31(11), 1751–1762. Retrieved from http://www.ncbi.nlm.nih.gov/pubmed/20162602

Napolitano, A., Ungania, S., & Cannat, V. (2012). Fractal Dimension Estimation Methods for Biomedical Images. In MATLAB - A Fundamental Tool for Scientific Computing and Engineering Applications - Volume 3. https://doi.org/10.5772/48760

Pfefferbaum, A., Rohlfing, T., Rosenbloom, M. J., & Sullivan, E. V. (2012). Combining atlas-based parcellation of regional brain data acquired across scanners at 1.5T and 3.0T field strengths. NeuroImage. https://doi.org/10.1016/j.neuroimage.2012.01.092

Pierpaoli, C., & Basser, P. J. (1996). Toward a quantitative assessment of diffusion anisotropy. Magnetic Resonance in Medicine, 36(6), 893–906. Retrieved from http://www.ncbi.nlm.nih.gov/pubmed/8946355

Pinto, M. S., Paolella, R., Billiet, T., Van Dyck, P., Guns, P. J., Jeurissen, B., … Sijbers, J. (2020). Harmonization of Brain Diffusion MRI: Concepts and Methods. Frontiers in Neuroscience, https://doi.org/10.3389/fnins.2020.00396

Resnick, S. M., Goldszal, A. F., Davatzikos, C., Golski, S., Kraut, M. A., Metter, E. J., … Zonderman, A. B. (2000). One-year age changes in MRI brain volumes in older adults. Cerebral Cortex, https://doi.org/10.1093/cercor/10.5.464

Reuter, M., Schmansky, N. J., Rosas, H. D., & Fischl, B. (2012). Within-subject template estimation for unbiased longitudinal image analysis. NeuroImage, 61(4), 1402–1418. https://doi.org/10.1016/j.neuroimage.2012.02.084

Ritchie, S. J., Bastin, M. E., Tucker-Drob, E. M., Maniega, S. M., Engelhardt, L. E., Cox, S. R., … Deary, I. J. (2015). Coupled changes in brain white matter microstructure and fluid intelligence in later life. Journal of Neuroscience, https://doi.org/10.1523/JNEUROSCI.0862-15.2015

Roberts, J. A., Perry, A., Roberts, G., Mitchell, P. B., & Breakspear, M. (2017). Consistency-based thresholding of the human connectome. NeuroImage. https://doi.org/10.1016/j.neuroimage.2016.09.053

Rubinov, M., & Sporns, O. (2010). Complex network measures of brain connectivity: uses and interpretations. NeuroImage, 52(3), 1059–1069. Retrieved from http://www.ncbi.nlm.nih.gov/pubmed/19819337

Schmitz, B. L., Aschoff, A. J., Hoffmann, M. H. K., & Grön, G. (2005). Advantages and pitfalls in 3T MR brain imaging: a pictorial review. American Journal of Neuroradiology, 26(9), 2229–2237.

Schönbrodt, F. D., & Perugini, M. (2013). At what sample size do correlations stabilize? Journal of Research in Personality. https://doi.org/10.1016/j.jrp.2013.05.009

Shrout, P. E., & Fleiss, J. L. (1979). Intraclass correlations: uses in assessing rater reliability. Psychological Bulletin, 86(2), 420–428. Retrieved from http://www.ncbi.nlm.nih.gov/pubmed/18839484

Smith, S. M. (2002). Fast robust automated brain extraction. Human Brain Mapping, 17(3), 143–155. https://doi.org/10.1002/hbm.10062

Smith, S. M., Jenkinson, M., Woolrich, M. W., Beckmann, C. F., Behrens, T. E. J., Johansen-Berg, H., … Matthews, P. M. (2004). Advances in functional and structural MR image analysis and implementation as FSL. In NeuroImage, https://doi.org/10.1016/j.neuroimage.2004.07.051

Srinivasan, D., Erus, G., Doshi, J., Wolk, D. A., Shou, H., Habes, M., & Davatzikos, C. (2020). A comparison of Freesurfer and multi-atlas MUSE for brain anatomy segmentation: Findings about size and age bias, and inter-scanner stability in multi-site aging studies. NeuroImage. https://doi.org/10.1016/j.neuroimage.2020.117248

Tanenbaum, L. N. (2005). 3T MRI in clinical practice. Applied Radiology.

Tardif, C. L., Collins, D. L., & Pike, G. B. (2010). Regional impact of field strength on voxel-based morphometry results. Human Brain Mapping, https://doi.org/10.1002/hbm.20908

Tax, C. M., Grussu, F., Kaden, E., Ning, L., Rudrapatna, U., John Evans, C., … Veraart, J. (2019). Cross-scanner and cross-protocol diffusion MRI data harmonisation: A benchmark database and evaluation of algorithms. NeuroImage. https://doi.org/10.1016/j.neuroimage.2019.01.077

Taylor, A. M., Pattie, A., & Deary, I. J. (2018). Cohort profile update: The Lothian birth cohorts of 1921 and 1936. International Journal of Epidemiology, https://doi.org/10.1093/ije/dyy022

Tustison, N. J., Cook, P. A., Klein, A., Song, G., Das, S. R., Duda, J. T., … Avants, B. B. (2014). Large-scale evaluation of ANTs and FreeSurfer cortical thickness measurements. NeuroImage. https://doi.org/10.1016/j.neuroimage.2014.05.044

Van Den Heuvel, M. P., Scholtens, L. H., Van Der Burgh, H. K., Agosta, F., Alloza, C., Arango, C., … De Lange, S. C. (2019). 10kin1day: A bottom-up neuroimaging initiative. Frontiers in Neurology, https://doi.org/10.3389/fneur.2019.00425

Wardlaw, J. M., Bastin, M. E., Valdés Hernández, M. C., Maniega, S. M., Royle, N. A., Morris, Z., … Deary, I. J. (2011). Brain aging, cognition in youth and old age and vascular disease in the Lothian Birth Cohort 1936: Rationale, design and methodology of the imaging protocol. International Journal of Stroke, https://doi.org/10.1111/j.1747-4949.2011.00683.x

Wardlaw, J. M., Brindle, W., Casado, A. M., Shuler, K., Henderson, M., Thomas, B., … Hernandez, M. C. V. (2012). A systematic review of the utility of 1.5 versus 3 Tesla magnetic resonance brain imaging in clinical practice and research. European Radiology, https://doi.org/10.1007/s00330-012-2500-8

West, J., Blystad, I., Engström, M., Warntjes, J. B. M., & Lundberg, P. (2013). Application of Quantitative MRI for Brain Tissue Segmentation at 1.5 T and 3.0 T Field Strengths. PLoS ONE, https://doi.org/10.1371/journal.pone.0074795

Wonderlick, J. S., Ziegler, D. A., Hosseini-Varnamkhasti, P., Locascio, J. J., Bakkour, A., Van Der Kouwe, A., … Dickerson, B. C. (2009). Reliability of MRI-derived cortical and subcortical morphometric measures: effects of pulse sequence, voxel geometry, and parallel imaging. NeuroImage, 44(4), 1324–1333. Retrieved from http://www.pubmedcentral.nih.gov/articlerender.fcgi?artid=2739882&tool=pmcentrez&rendertype=abstract

Yamada, H., Abe, O., Shizukuishi, T., Kikuta, J., Shinozaki, T., Dezawa, K., … Imamura, Y. (2014). Efficacy of distortion correction on diffusion imaging: Comparison of FSL eddy and eddy-correct using 30 and 60 directions diffusion encoding. PLoS ONE, https://doi.org/10.1371/journal.pone.0112411

Zhang, H., Schneider, T., Wheeler-Kingshott, C. A., & Alexander, D. C. (2012). NODDI: Practical in vivo neurite orientation dispersion and density imaging of the human brain. NeuroImage. https://doi.org/10.1016/j.neuroimage.2012.03.072

